# Strict lockdown versus flexible social distance strategy for COVID-19 disease: a cost-effectiveness analysis

**DOI:** 10.1101/2020.09.14.20194605

**Authors:** Ben W. Mol, Jonathan Karnon

## Abstract

**Objectives:** To balance the costs and effects comparing a strict lockdown versus a flexible social distancing strategy for societies affected by Coronavirus-19 Disease (COVID-19).

**Design:** Cost-effectiveness analysis.

**Participants:** We used societal data and COVID-19 mortality rates from the public domain.

**Interventions:** The intervention was a strict lockdown strategy that has been followed by Denmark. Reference strategy was flexible social distancing policy as was applied by Sweden. We derived mortality rates from COVID-19 national statistics, assumed the expected life years lost from each COVID-19 death to be 11 years and calculated lost life years until 31^st^ August 2020. Expected economic costs were derived from gross domestic productivity (GDP) statistics from each country’s official statistics bureau and forecasted GDP. The incremental financial costs of the strict lockdown were calculated by comparing Sweden with Denmark using externally available market information. Calculations were projected per one million inhabitants. In sensitivity analyses we varied the total cost of the lockdown (range −50% to +100%).

**Main outcome measure:** Financial costs per life years saved.

**Results:** In Sweden, the number of people who died with COVID-19 was 577 per million inhabitants, resulting in an estimated 6,350 life years lost per million inhabitants. In Denmark, where a strict lockdown strategy was installed for months, the number of people dying with COVID-19 was on average 111 per million, resulting in an estimated 1,216 life years per million inhabitants lost. The incremental costs of strict lockdown to save one life year was US$ 137,285, and higher in most of the sensitivity analyses.

**Conclusions:** Comparisons of public health interventions for COVID-19 should take into account life years saved and not only lost lives. Strict lockdown costs more than US$ 130,000 per life year saved. As our all our assumptions were in favour of strict lockdown, a flexible social distancing policy in response to COVID19 is defendable.

## Introduction

The Coronavirus disease 2019 (COVID-19) pandemic caused by the SARS-CoV-2 virus (Corona-virus) has disrupted societies around the world.^1^ At present (Early September 2020), around the world more than 27 million people have been diagnosed with the virus and almost 1,000,000 of them have died.

The consequences of the response to COVID-19 are unprecedented. Economies have crashed leaving hundreds of millions unemployed, with governments reserving trillions of dollars for immediate health care, to save large and small businesses and support affected individuals. Unsurprisingly, there is been fierce debate on the optimal public health policy in response to COVID-19. Among the many problems that COVID-19 brings, there are two main issues. First, since patients with COVID-19 pneumonia require often acute respiratory support, there is a threat of overload of medical systems, specifically intensive care capacity. Without public health response, exponential spread of the virus will stress intensive care capacity, as has been seen in Wuhan, Lombardy, Madrid, Strasbourg, London and New York. Second, COVID-19 has a considerable mortality in individuals > 65 year and individuals with co-morbidity or obesity, while mortality in healthy individuals < 65 years is relatively low.^2^

To address these problems, all countries confronted with COVID-19 have put public health strategies in place, varying from social distancing recommendations to nationwide lockdowns, commonly including for example stay-at-home orders, school closures, curfew and restrictions on within-country travel. The aim of these strategies is either suppression - reverse epidemic growth, reducing case numbers to low levels and maintaining that situation indefinitely (while waiting for a vaccine or better treatment options) – or mitigation (flatten the curve), the latter aiming to slow but not necessarily stop epidemic spread.^3,4^ Herd immunity might be a side product of mitigation.^5^

While many involved countries have implemented a strict lockdown, Sweden went against the grain by keeping public life less restricted.^6^ Here, we perform an economic analysis balancing costs and effects of a strict lockdown versus a more liberal social distancing strategy by comparing Sweden to Denmark.

## Methods

We performed a cost-effectiveness analysis in which we compared the costs and effects of a strict lockdown versus a more liberal social distancing strategy, the latter being the reference strategy. For the strict lockdown strategy, we used data from Denmark. For the liberal social distancing strategy, we took data from Sweden. Calculations were adjusted for differences in population size, and expressed per million inhabitants. Our effectiveness measure was the number of life years saved, while the time horizon was 6 months.

### Data sources

For both countries, we extracted factual information on COVID-19 mortality rates from the most up-to-date situational reports over the period 1^st^ March – 31^st^ August 2020.^7 8^ Information was extracted independently in duplicate by two authors (BW, JK) and discrepancies were resolved. These data provided the total number of people who died from COVID-19 between March 2020 until August 31^st^ 2020.^9^

We assumed that the mean number of lost life years of someone dying with COVID-19 was 11 years.^10 11^ Medical costs of COVID-19 were only counted from hospital admissions. The costs of one hospital admission for COVID-19 disease were estimated to be US$ 8,400,^12^ and for each person dying with COVID-19 we assumed 9 hospital admissions for non-fatal COVID cases, which compares to the average case-fatality rate of COVID-19 in the Scandinavian countries, while in a Chinese study the case-fatality rate among hospitalized patients was higher.^13^

The expected economic costs were derived from GDP statistics from each Nordic country’s official statistics bureau and forecasted GDP based on the SEB Nordic Outlook 2020 (released August 25^th^ 2020).^14^ Sweden’s forecasted GDP decline in 2020 is estimated at 3.8%. We compared the forecast for Sweden with other current projections (Handelsbanken, Danske Bank, Swedbank, the IMF, The National Institute of Economic Research (Sw: ‘Konjunkturinstitutet’) and the Swedish Government and note that projections range between negative 4-7% for 2020, with most estimates ranging between negative 6.5-7.0%. As such, we have deemed the SEB projections to be reasonable and used them as a basis for the analysis. From this analysis, the cost for implementing a strict lockdown of the economy (closing of hotels, restaurants, shops, travel etc.) is unsurprisingly higher per million inhabitants in Denmark (US$ 2.7bn) compared to Sweden (US$ 1.9bn). We then adjusted the GDP loss in Denmark for the difference in GDP per million as compared to Sweden.

For indicative purposes, we have applied the year-to-year growth projections per country on reported 2019 GDP to arrive at a 2020 GDP figure (for Sweden these data were not available, thus we multiplied reported 2018 GDP to a projected growth of 1.1% for 2019, and subsequently that figure multiplied by the projected negative 6.5%). In the next stage, we have translated the local currencies to US$ using the 2019 average exchange rate. We further note that our current projections hinges on a recovery of GDP in Q3-Q4 2020 from an incremental opening of the economies around the world. Should the current situation not be resolved or at least eased in the near term, then our projections appear optimistic. This is addressed in a sensitivity analysis in which the relative increase in costs in Denmark was varied between −50% and +100%, respectively. For the lockdown strategy, the projected economic costs were made under the assumption that there would be an ease in the lockdown after two-three months. As our time horizon was 6 months in the baseline case, we did not discount costs.

### Description of the strategies

The <u>strict lockdown strategy</u> advised people to stay at home, limited movements of a community to necessary travel for food, medical need and travel to and from work that was deemed to be necessary for society. Work should be done from home if possible, and schools, universities, hotels, cafés, and restaurants were closed. Gathering of more than 10 people was not allowed, with the exemption of people living in the same household. Elective medical procedures were stopped. Such measured were put in place in Denmark mid-March 2020, and were eased after two to three months, i.e. from mid-May onwards.

The <u>more liberal social distancing strategy</u> allowed people to go out. Schools (scholars < 16 years), restaurants and shops stayed open, and people were allowed to go to work. Gatherings of up to a group size of 50 people were allowed. There were no restrictions on within-country travel, but there was an advice against unnecessary travel. Social distancing was encouraged by the government, leaving a large amount of self-responsibility with individual members of society. The medical system continued to operate, albeit with a strong reduction in elective interventions, although preparations were made to deal with an increased number of patients with COVID-19. Details on the strategies can be read at http://www.oecd.org/coronavirus/en/.

### Cost-effectiveness analysis

We compare the costs and effects of lockdown versus a more liberal social distancing strategy policy. Both interventions, the strict lockdown strategy and the more liberal social distancing strategy were implemented mid-March 2020. By August 2020, after Denmark had ended its lockdown, COVID-19 policies between the countries were comparable, and very much followed the more liberal social distancing strategy that Sweden had applied initially. We estimated for each country (per million inhabitants) the expected total number of life years lost from COVID-19 by 31^st^ August 2020 by multiplying the number of persons who died by 31^st^ August 2020 with COVID-19 with a mean life expectancy of 11 years.^10^ As statistics do not allow distinction between persons who die from COVID-19 or persons who die from other causes while being infected with Corona-virus, we refer to anyone dying while infected with Corona-virus as a COVID-19 related death, independent of the cause.

The incremental effectiveness of a strict lockdown policy over a more liberal social distancing strategy was then obtained by subtracting the expected number of life years lost in Denmark from the expected number of life years lost in Sweden. In the initial analysis, we used the weighted average of Denmark, while the results were also calculated for each of these countries separately. The incremental costs were calculated from the difference between the cost (per million inhabitants) in Sweden and each of the other countries.

The incremental cost-effectiveness ratio (ICER) was calculated from the ratio of the incremental expected costs and estimated incremental life years saved between each of the other countries and Sweden.

### Sensitivity analyses

In sensitivity analyses, we varied lost life expectancy of patients dying with COVID-19 (base case 11 years, range 1-11 years), and cost of implementing strict lockdown with −25%, +25%, +50% and +100.

## Results

Table 2 shows the main results. Sweden applied since early March 2020 the more liberal social distancing strategy. Denmark introduced strict lockdowns on 13 March 2020, respectively. In Sweden, the number of people dying with COVID-19 per million inhabitants acquired by August 31^st^ was 577, resulting in 6,350 life years lost per million inhabitants. In Denmark the number of people dying with COVID-19 was 111 per million, resulting in 1,216 life years lost. Compared to Sweden, the Lockdown in Denmark resulted per million inhabitants in 466 lives saved and 5,134 life years gained as compared to Sweden by August 31^st^.

**Table 1:** Baseline assumptions

|  | Baseline value | Range<br>(sensitivity analysis) |
| --- | --- | --- |
| Expected lost live years from one COVID-19 death | 11 years | 2 to 15 years |
| Costs of one hospital admission for COVID-19 | US\$ 8,400 | futile |
| Mortality risk of patients with COVID-19 admitted to hospital | 10% | futile |
| Loss in life expectancy from reduction in elective regular health care /screening | Not considered |  |
| Loss in life expectancy from reduced future health care due to loss of health insurance or savings in the public health system | Not considered |  |
| Cost of implementing strict lockdown in US\$ (per million inhabitants) | Denmark US\$2.7 bn | -50%, -25%, +25%,<br>+50% and +100% |
| Cost of implementing flexible social distancing in US\$ (per million inhabitants) | Sweden US\$1.9 bn | |

**Table 2:** Main results

|  | Denmark (DK) | Sweden (S) |
| --- | --- | --- |
| Population (million) | 5.8 | 10.1 |
| First day with > 100 cases diagnosed | 10-mrt. | 6-mrt. |
| First day with > 10deaths | 23-mrt. | 18-mrt. |
| Absolute Number of people positive for corona-virus | 16985 | 45942 |
| Number of people positive for corona-virus / million inhabitants | 2928 | 8356 |
| Number of people dying with corona-virus | 641 | 5830 |
| Number of patients dying with corona-virus / million inhabitants | 111 | 577 |
| Mean live expectancy (years) | 11 | 11 |
| Expected deaths from Corona (million inhabitants) | 111 | 577 |
| Expected life years lost from Corona (million inhabitants) | 1216 | 6350 |
| Gained lives / million inhabitants | 467 | reference |
| Gained live years / million inhabitants | 5134 | reference |
| Value of 1 life-year (US\$) | 100,000 | 100,000 |
| Justifiable cost /million inhabitants (threshold \$ 100,000 /life year saved) (bill US\$) | 0.51 | reference |
| Justifiable cost / for threshold \$ 100,000 /life year saved) (billion US\$ ) | 3.0 | reference |
| Medical cost from COVID-19 per million inhabitants (million US\$) | 9.3 | 48 |
| Extra economic cost/ million inhabitants (million US\$) | \$744 | reference |
| Diff medical + government cost/ million inhabitants (million US\$) | \$705 | reference |
| **Cost per life year saved (US\$)** | **\$137,285** | **reference** |

For direct hospital care for COVID-19 patients for Sweden, the total costs are expected to be US$ 48.5 million per million inhabitants, while for Denmark these total costs were US$ 9.3 million per million inhabitants.

The additional costs were estimated by comparing the absolute decrease between reported 2019 GDP and projected 2020 GDP per country, converted with the yearly average 2019 US$ exchange rate. Per million inhabitants, a US$ 1.9 billion decrease (3.9%) per million inhabitants was estimated for Sweden. The additional costs for Denmark represent the variance to the Sweden estimate. The additional reduction in GDP per million inhabitants was estimated to be US$ 743 million per million inhabitants.

The incremental costs to save one life year from a strict lockdown were therefore US$ 137,285 per life year saved.

Threshold analysis indicated that for the base case, and assuming US$ 100,000 as maximum cost to be spend for a life year saved, Denmark would per million inhabitants be willing to spend US$ 510 million per million inhabitants. This corresponds with a total additional budget to be spent of US$ 3.0 billion.

In August, SARS-CoV-2 infection rates were low in all three countries, with 73 per million inhabitants in Sweden, while this rate was 9 per million inhabitants per month in Denmark. In August, COVID-19 related mortality rates were 7 and 2 per million inhabitants per month for Sweden and Denmark, respectively.

### Sensitivity analysis

Sensitivity analysis indicated that variation of the lost-life years from an individual dying with COVID-19 between 15 years and 2 years resulted in a cost-effectiveness varying between US$100,676 and US$755,068 per life year saved. Variation of the economic impact of Lockdown between −50% and +200% resulted in a cost-effectiveness varying between US$64,824 and US$282,207.

## Discussion

In this cost-effectiveness analysis, we compared a strict lockdown for COVID-19 disease as implemented in Denmark to the more liberal social distancing policy applied in Sweden. Under current Swedish policy 577 more people per million inhabitants died a COVID-19 related mortality as compared to Denmark, resulting in an additional 1,216 life years lost. The expected incremental costs that Denmark made to save one life year using Sweden as a reference was US$ 137,285. This finding was relatively robust in our sensitivity analyses. Our study has several strengths. Denmark and Sweden were, prior to the COVID-19 outbreak and the subsequent public health response, very comparable from a demographic perspective, and to a lesser extent also from an economic perspective.^15^ Our data on COVID-related mortality and reduction in GDP were obtained in an unbiased and similar way in each of the four countries. We confirmed the COVID-19 mortality with the national birth statistics. In Denmark the absolute number of people that died between March-August 2020 (week 10-34) was on average 64 per million higher than in 2015-2019, while in Sweden this number was 502 per million higher (difference 438 per million). In our calculations, the difference between Sweden and Denmark was very comparable with 503 COVID-19 related additional deaths per million in Sweden. Also, as there is considerable debate on the contagiousness and mortality of SARS-CoV-2, we only used COVID-19 related mortality rates per million and economic impact.

The profile of other Scandinavian countries might be comparable to Sweden and Denmark. From an economic perspective, Norway has a much higher GDP per capita than the other Scandinavian countries, partially related to its oil-industry. Therefore, a relatively high decrease in GPD will still resulted in a relatively small reduction in GDP after correction for the GDP per capita difference with Sweden, making the relative cost of the lockdown in Norway relatively low. Finland had a lowed reduction in GDP than Sweden, despite its Lockdown. The best comparison is therefore probably between Sweden and Denmark, clearly showing that Lockdown is not cost-effective.

We probably have underestimated the impact of the effect of the lockdown on GDP, as the expected reduction in the Swedish GDP is partly driven by a shrinkage in revenues of international companies, also affecting Sweden’s GDP.^16^ Closing of schools as part of the strict lockdown has a strong impact on costs, which was not specifically incorporated. A Norwegian study reported closing of schools to cost 170 million US$ per day, for a total of 8 billion until the rest of this school year, with similar calculations on the Sweden side.^17^ This means that closing of the schools alone already was more expensive than was allowed according our threshold analysis to make a strict lockdown cost-effective.

All our assumptions were also in favour of the lockdown policy. We expected 11 years of lost life expectancy from a person dying from COVID-19 of someone at the age of 79. However, as COVID-19 patients often have co-morbidity and/or obesity, their life expectancy is likely to be lower. We also did not take into account the loss in life expectancy from the reduction in elective regular health care and screening due to the lockdown.^18^ Of more concern is the long-term impact of the strict lockdown on health, which is also not incorporated in our analysis. Employment changes during and after the 2008 financial crisis had a strong adverse effect on chronic health for five broad types of health conditions, with the strongest effects being for mental health conditions, and this is likely to happen again.^19 20^

Also, we did not incorporate disability-adjusted life years, representing a weighted combination of mortality and morbidity effects of an intervention. On one hand, COVID-19 infection affects the quality of life of those who survive the infection.^21^ On the other hand suffer most dying from COVID-19 already disabilities, thus reducing the impact of COVID-19 related death.

Best ways out of the current crisis is either a vaccine against SARS-CoV-2 infection or better treatment of COVID-19. Recent randomised trials have indicated that antiviral treatments and dexamethasone both potentially improve time to recovery and decrease mortality in COVID-19 disease with about 30%.^22–24^ Such a decrease makes a strict lockdown 30% less cost-effective, as costs stay roughly the same but effectiveness decreases.

On the other side of the spectrum, if a vaccine were not to be available in the near future, then the less strict Swedish social distancing policy is likely to lead to herd immunity at some stage. With 7% of the Swedish population infected by early May 2020 and 60% needed for herd immunity, it will take under the current speed of spread of the infection another 15 months to achieve that. After that time, the infection rate will decrease in Sweden, whereas other countries either also have to accept new infections, or they have to reinforce the lock down. Both scenarios will make lockdown less cost-effective.

A less strict social distancing policy that accepts a slow spread of the infection requires good health. The health of the population in the Nordic countries, with obesity rates between 20-25% facilitates a liberal social distancing policy better than for example the United States, where rates of obesity and obesity related co-morbidity are much higher.^25^ Alternatively, public health interventions should consider advocating a healthy lifestyle. Even in the relatively healthy Nordic Countries, adhering to the fruits and vegetable intakes would increase the percentage of deaths delayed or saved by 30-50%, very comparable or even better than strict lockdown, and at a fraction of its cost.^26^

Our study does not want to convince for a policy pro or contra strict lockdown. In view of the extreme impact that COVID-19 disease has on health and health systems when it is left uncontrolled, a first response of lockdown is understandable and defendable. However, our analysis also shows that the Swedish more liberal policy, despite it higher mortality rates, is acceptable and in terms of cost-effectiveness not as bad as is perceived when one assesses the absolute number of COVID-19 related deaths.

The National Institute for Health and Care Excellence (NICE) in the United Kingdom adopts cost-per-lifeyear saved threshold of US 30,000 per life year saved.^27^ In 2016, NICE set the cost-per-lifeyear threshold at US$ 100,000 for treatments for rare conditions because, otherwise, drugs for a small number of patients would not be profitable.^28^[3] Thus, the US$ 137,285 per life year saved that we calculated indicates that the policy of strict Lockdown is not indisputable.

As such, we want to suggest changing the metrics that governments and policy makers are using to lost life-years, rather than absolute numbers. Also, while many societies are releasing strict lockdown programs, renewed infection should not lead to panic and radical renewed lockdown. It is important that we look critically at each of the individual lockdown measures, and understand in which conditions SARS-CoV-2 spreads. Similarly, Sweden will learn from situations where SARS-CoV-2 infections resulted in high mortality, for example nursing homes. Stratified shielding, in which high risk groups are protected while spread of the infection is accepted, is a good compromise.^29^

## Data Availability

We used societal data and COVID-19 mortality rates from the public domain.

https://www.worldometers.info/coronavirus/

https://news.google.com/covid19/map?hl=en-AU&gl=AU&ceid=AU:en

## Acknowledgement

The authors thank Corine Verhoeven, Janneke van ‘t Hooft and Cathrine Axfords for critically reviewing the manuscript, and Hildegard Mostmans for her help with referencing and lay-out.

